# Coping with drug resistant tuberculosis alongside COVID-19 and other stressors in Zimbabwe: a qualitative study

**DOI:** 10.1101/2023.02.24.23286187

**Authors:** Collins Timire, Katharina Kranzer, Debora Pedrazzoli, Fungai Kavenga, Samuel Kasozi, Fredrick Mbiba, Virginia Bond

## Abstract

**Background:** Households in low-resource settings are more vulnerable to events which adversely affect their livelihoods, including shocks such as the death of a family member, inflation, droughts and more recently COVID-19. Drug Resistant Tuberculosis (DR-TB) is also another shock that inflicts physical, psychological and socioeconomic burden on individuals and households. We describe experiences and coping strategies among people affected by DR-TB and their households in Zimbabwe during the COVID-19 pandemic, 2020 to 2021.

**Methods:** We conducted 16 in-depth interviews with adults who had just completed or were completing treatment. Interview themes included health seeking behaviour, impact of DR-TB on livelihoods and coping strategies adopted during treatment. We analysed data using thematic analyses.

**Results:** Health seeking from providers outside the public sector, extra-pulmonary TB and health system factors resulted in delayed DR-TB diagnosis and treatment and increased financial drain on households. DR-TB reduced productive capacity and narrowed job opportunities leading to income loss that continued even after completion of treatment. Household livelihood was further adversely affected by lockdowns due to COVID-19, outbreaks of bird flu and cattle disease. Stockouts of DR-TB medicines, common during COVID-19, exacerbated loss of productive time and transport costs as medication had to be accessed from other clinics that were further away. Reversible coping strategies included: reducing number of meals; relocating in search of caregivers and/or family support; spending savings; negotiating with school authorities to keep children in school. Some households had to adopt irreversible coping strategies such as selling productive assets and withdrawing children from school.

**Conclusion:** DR-TB combined with COVID-19 and other stressors pushed households into deeper poverty, and vulnerability. Multi-sectoral approaches that combine health systems, psychosocial and economic interventions are crucial to mitigate diagnostic delays and suffering, and meaningfully support people with DR-TB and their households to compensate the loss of livelihoods during and post DR-TB treatment.

## Background

Tuberculosis (TB) is a global public health challenge, and until the COVID-19 pandemic, was the leading cause of death from a single infectious disease (ranking above HIV).(1) An estimated 10.6 million people fell ill with TB in 2021 and 1.6 million (15%) of them died.(1) Treatment success (cure and treatment completion) for people with drug resistant TB (DR-TB) is worse, (averaging around 60% globally and around 54% in Zimbabwe)(2–4), compared to 86% for people with drug-susceptible TB (DS-TB).(1,5) People with DR-TB have higher risk of loss to follow up, death and post-TB disabilities including poor lung function, auditory complications and, depending on site of infection, bone deformities.(6–8)

Successful treatment outcomes among people with DR-TB depend on completing appropriate and efficacious course of medicines. Until very recently, treatment of DR-TB was much longer and more toxic than that of drug susceptible TB (DS-TB), lasting up to 18-24 months. As a result, people affected by DR-TB and their households often have to overcome even greater health system, psychosocial and socioeconomic barriers over an extended time before completing treatment.(9–12) Surveys of cost incurred by TB patients and their households have shown that a higher proportion of people affected by DR-TB incur catastrophic costs than those affected by DS-TB (82% vs 47%).(13–15) While the greater proportion of TB related costs are indirect costs such as income loss due to reduced productivity and time spent health seeking,(16) people and their households also incur out-of-pocket costs which may be substantial to poor people who are less likely to have medical insurance, savings or stable earnings.(17) Health system approaches that have been implemented globally to reduce TB related costs include: i) early diagnosis of TB through systematic screening and universal access to drug susceptibility testing (18), ii) decentralisation of TB treatment to reduce transport costs, iii) provision of free TB medication and diagnostics and iv) provision of psycho-social and economic support. Notwithstanding these interventions, costs of hospitalisation, radiology services (chest X-rays), ancillary medicines, consultations and blood tests are often not considered, (19) and individuals and their primary caregivers also incur substantial income loss (or indirect costs) related to taking time off work to seek and receive TB care.(20,21) Such indirect costs, together with costs related to food and transport to/from the health facility, constitute the major cost drivers.(19–21)

In addition to economic losses, households also experience worsening social relations and stigma.(24) Further shocks and stressors may affect households either concurrently or sequentially.(25) Such shocks may be spatial or covariate (e.g., COVID-19, drought, floods) when they affect communities, nations, regions or the whole world or may be temporal based on their occurrence (annual, seasonal).(26) Often shocks have synergistic relationships that determine household coping strategies.(25) Poor and distressed households ― usually without medical insurance or savings ― may adopt reversible coping strategies such as delaying health seeking for chronic diseases, relocating in search of food or caregivers and spending savings, and faced with acute infections or global shocks, may be forced to mobilise and expend their resources rapidly.(27) Under such circumstances, they may be forced to adopt irreversible coping strategies such as taking loans at exorbitant interests, withdrawing children from school and selling productive assets, thereby deepening their vulnerability to current and future shocks.(27,28) COVID-19 is a global stressor and has been shown to be a stress multiplier.(29) Poor households that experience a convergence of DR-TB, COVID-19 and other stressors may have both a limited choice of coping strategies and limited coping capacity.(30)

Zimbabwe, a resource limited country in sub Saharan Africa wracked with economic hardship since 2000, had an incidence of TB and DR-TB of 190 per 100,000 population and 4.9 per 100,000 population, respectively, in 2021, and TB/HIV co-infection was 50%.(5,31,32) Around 90% of the population is unemployed and earn a living through informal jobs (subsistence farming, informal businesses as vendors and cross border trading), and 72% live in poverty.(33) From March 2020-August 2020, the Zimbabwean government imposed a stringent lockdown with the aim to control the spread of COVID-19. During this time, businesses were shut and public transport almost completely ceased. Poor people, especially those in urban areas, faced an increased risk of contracting COVID-19 owing to overcrowded living conditions (34) and bore the brunt of lockdowns since they mostly live from daily earnings and are informally employed with no social security. In the absence of social support, COVID-19 thus negatively affected household income and food security. It is well established that poverty fuels both COVID-19 and TB and in turn, the two diseases deepen impoverishment and irreversible coping strategies.(27,30) Previous studies have looked at the impact of DR-TB, but not in the context of global shocks such as COVID-19.(9,35,36) COVID-19 affected whole populations and impacted the health systems, livelihood and transportation. To study the joint impact of DR-TB, COVID-19 and other stressors, we conducted 16 in-depth interviews with people affected by DR-TB in Zimbabwe in 2020-2021 to trace their experiences and coping strategies during the pandemic and their period of treatment.

## Methods

### Management of DR-TB in Zimbabwe

In Zimbabwe, DR-TB care is provided at primary health care level. Prior to 2017, treatment duration was 18-24 months with daily injections for the first six months. From 2017, shorter (9-12 months) injection-based regimens were introduced followed by the introduction of a short all oral 9 months regimen in 2020.(37) Xpert MTB/Rif (Cepheid, Sunnyvale, CA, USA) is the primary diagnostic test for people presenting with signs and symptoms of TB enabling not just diagnosis of TB, but also detection of rifampicin resistance.(37) People on DR-TB treatment are eligible for USD 25 conditional cash transfers and an additional USD 20 worth of grocery vouchers (from 2021 onwards) every month for the duration of treatment. However, delays and inconsistencies in cash disbursements and food hampers have been reported especially during the COVID-19 pandemic, with only two-thirds (64%) of those registered effectively receiving cash.(38)

### Study design, population and recruitment

This was a qualitative phenomenological study to gather lived experiences of people with DR-TB. Men and women aged ≥18 years who were within two months of completing DR-TB treatment or had completed treatment within two months were identified from TB registers within health facilities in Harare (urban) and Matebeleland South (rural) provinces in Zimbabwe. The study team met or called the potential participant and appraised them about the study. Those who expressed willingness to participate in the study were asked to identify a convenient place and time so that the study team could obtain written informed consent and conduct in-depth interviews.

### Data collection

In-depth interviews were conducted from October 2020 to March 2021 by CT (first author) and FM (social scientist), with CT conducting 12 interviews and FM four interviews. The interview guide, translated into two local languages (Shona and Ndebele), included the following themes: health seeking patterns, experiences with DR-TB treatment and DR-TB disease, and coping strategies adopted during treatment. The interviews lasted 35-45 minutes and were conducted in Shona or Ndebele. All interviews were audio-recorded and were conducted in private locations suggested by participants. Participants received USD3 for refreshments, and bus fares were also reimbursed. After each interview, the two researchers (CT and FM) had debriefing meetings to discuss the findings.

### Ethical approval

Ethical approval was obtained from the London School of Hygiene and Tropical Medicine Research Ethics Committee (Ref 22579), the Biomedical Research and Training Institute Institutional Review Board (AP160/2020) and the Medical Research Council of Zimbabwe (MRCZ/A/2645). Permission to access DR-TB registers was obtained from the Permanent Secretary for Health, Zimbabwe. All participants gave written informed consent to take part in the study and for the use of an audio-recorder. Names used in quotes and tables in this paper are pseudonyms.

### Data analysis

All audio-recordings were transcribed into local language before translating the transcripts into English. Data were analyzed using thematic analysis. First, the transcripts were read several times aiming to get familiar with the data. Second, data were coded manually and using *NVivo* version 12 software (QSR International). The codes were both data driven and concept driven, (39,40) and were grouped into themes to form thematic maps.(41) Divergent views were noted.

## Results

Sixteen people (eight women) were enrolled. Twelve were co-infected with HIV. Their characteristics are summarized in **Table 1**. We first describe the experiences of people with DR-TB starting from delays in treatment initiation (due to underdiagnoses, misdiagnosis of TB or health seeking from alternative providers), physical impact of DR-TB disease and treatment itself, and changes in livelihoods as a result of DR-TB episodes. Next we explore the coping strategies adopted in response to DR-TB alongside other shocks, including COVID-19.

**Table 1:** Characteristics of DR-TB patients who were enrolled in the study, Zimbabwe, 2020-2021

| Identifier | HIV status | Regimen | Marital status | History of TB | Employed | Site of TB | Residency | Received DR-TB allowance | Stage of interview |
| --- | --- | --- | --- | --- | --- | --- | --- | --- | --- |
| Agnella_F‡ | Positive | 18-mo | Single mother of 2 children | No | No | PTB | Urban | Yes§ | Post-treatment |
| Ruth_F | Positive | 9-11mo | Single mother | Yes | No | PTB | Urban | Yes§ | Post-treatment |
| Felistus_F | Negative | 18-mo | Single, never married | Yes | No | PTB | Urban | Yes§ | During treatment |
| Moses_M | Negative | 9-11mo | Married | No | No | PTB | Urban | Yes§ | During treatment |
| Peter_M | Positive | 18-mo | Single, never married | No | No | PTB | Urban | No | Post-treatment |
| Caleb_M | Positive | 9-11mo | Married | No | No | PTB | Rural | Yes § | During treatment |
| Talent_F | Positive | 18-mo | Single mother | No | No | EPTB | Urban | No | Post-treatment |
| Lydia_F | Positive | 18-mo | Single, never married | No | Yes | PTB | Rural | Yes within 3 months | During treatment |
| Godfrey_M | Positive | 18-mo | Single, never married | Yes | No | PTB | Rural | No | Post-treatment |
| Sipho_M | Positive | 18-mo | Married | Yes | No | PTB | Rural | Yes§ | During treatment |
| Spiwe_F | Negative | 9-11mo | Married | No | No | PTB | Rural | No | During treatment |
| Loveness_F | Negative | 18-mo | Single mother (1child) | No | No | EPTB | Urban | Yes within 3 months | Post-treatment |
| Tecla_F | Positive | 18-mo | Single mother (1child) | No | No | EPTB | Urban | Yes within 3 months | Post-treatment |
| Nyenge_M | Positive | 9-11mo | Single | Yes | No | PTB | Rural | Yes within 3 months | During treatment |
| Wembe_M | Positive | 9-11mo | Married | New Patient | No | EPTB | Rural | No | Post-treatment |
| Fanuel_M | Positive | 9-11mo | Married with 2 children | New patient | No | PTB | Rural | Yes within 3 months | During treatment |

*‡=Pseudonym and age; PTB=Pulmonary TB; EPTB=extra-pulmonary TB; § First cash disbursement came after at least 8 months*

**Table 2:** Additional quotes and themes

| *Themes* | *Subtheme* | *Quotes supporting themes* |
| --- | --- | --- |
| *Health seeking* | *Alternative providers* | *We were also advised by the traditional healers that we are supposed to go and consult hospitals and other traditional healers who help.....they mostly advised us to go to hospitals. That's when we started to go to Y [a private clinic]. Caleb_M* |
|  |  | *...we consulted another herbalist and he informed us that there was need for two white goats with which he performed the rituals...He charged us R2500 (USD 170) but my father negotiated with him. He gave him R1000 (USD 70) and promised to bring the balance of R1000. Caleb_M* |
|  |  | *On the first days when I was able to walk but was still weak, we visited a certain traditional healer. Caleb_M* |
|  | *Recognition of TB symptoms* | *On that day I didn't even drink alcohol. I tried but I couldn't. I couldn't even smoke. What is happening here? Could it be that thing again? It's that thing again! I was certain about that. I decided to wait for some change. If I didn't see any change I would go back to the hospital. I realized there was no change. Then I went back [to the hospital]. Godfrey_M* |
| *Health system constraints* | *Misdiagnosis and diagnostic delays* | *On that day they called and advised that my treatment be changed. That's how I was diagnosed with MDR-TB. So they spent a few days hesitating to tell me that I was here to stay. This was no longer time for jokes. I had to forget about the previous two months of admission. It was a joke [laughs]. I was going to be admitted for six months because of DR-TB. Godfrey_M* |
|  |  | *When I had completed treatment, they told me that I was supposed to be injected. Ruth_F* |
|  | *Long waiting times* | *So they isolated us. After they have finished whatever they would be doing, that's when they would come to the back to attend to us. After that we would go back home. Ruth_F* |
|  |  | *Those with common TB were served first but initially the sister-in-charge had told us that we were to be served first. But some of the times we had to wait for others to finish first....to wait for about 45 minutes to an hour then they would come and administer our injections. Then we would leave. Peter_M* |
|  | *Lack of privacy* | *...they would inject us while we were outside where we could be seen by people who stay close to us. There are some houses which do not have dura-walls, some are fenced...people would watch as we received our injections. Loveness_F* |
|  | *Delays in laboratory results* | *I am confused because if I give them a sputum sample so that my treatment is changed and I do not get the results it means I stay on the same treatment. Spiwe F* |
|  | *Drug stock-outs* | *.... they would ask us to go to X [clinic] and other poly clinics like Y. They would say, "We have run out of a certain drug." Then we would rush to either Z or to Y to look for it. This happened several times...Agnella_F.* |
| *Psychosocial, physical and economic impacts* | *Separation from partner* | *It affected me very much because it affected my marriage. The moment I got unwell, my husband could not accept it. He lacked the will to help me. I just don't know why. Maybe he thought I was going to die and he had nothing to do with me. Because the moment I got sick he ran away and went to South Africa. He never came back. Ruth_F* |
|  | *Hospitalisation and huge medical bills* | *Yes on the first instance I was admitted for 10 days and we paid the required fee. On the second occasion money was needed for a blood transfusion that's when I was admitted for 3 days whilst we were waiting for the blood. So when the blood was delivered a transfusion was conducted and we paid for the blood. Then on the third occasion I was admitted for 5 days because my kidneys were no longer functioning properly and we paid for the admission and the medicines. Tecla_F* |
|  | *Side effects of TB treatment* | *I managed to finish my medication even though I have some side effects, I experienced lots of side effects on my legs...they are numb such that I can't walk, for me to walk long distances. Agnella_F.* |
|  |  | *Like now, I have problem with my legs. The toes shiver on their own. You see that? I don't know what's causing the problem. I also have numbness from here [knees], and also in my joints and hands. I am supposed to consult a doctor or clinic to get help but I don't have the money. I need medical attention and food. Loveness_F* |
|  | *Narrowed job opportunities* | *I was once employed by company X where they collect garbage bins. The work environment has a lot of dust. The moment I got there, it didn't take me a long time before I got sick. I think I only worked for just a week and started coughing again...I later on quit the job. So this sickness affected me a lot. I can no longer get employed to do every other job except only those that pose less risk to my health. Ruth_F* |
|  |  | *I used to work as a DJ but ever since I was diagnosed with TB last year, I have never worked. In addition, I was advised that I must not mix and mingle with people in dusty environments before I finish my treatment since I might meet a TB infected person and may get infected. Caleb_M* |
| *Coping strategies* | *Selling assets* | *I sold them for 700 each and got R1,400 for both speakers. I bought them for R3,500 each meaning R7, 000 for the two. I was looking forward to sell them for R2, 000 each and would get R4, 000 for the 2 speakers. So, I sold them for R700 each because things were tough for us. Caleb_M* |
|  |  | *I sold 2 speakers. The old man [my father] sold a car. He sold his [Toyota] Wish. That time money was hard to come by. It took a long time before my wife got a kitchen [canteen] business. Caleb_M*<br>*We sold two goats. We sold them under pressure because I wanted to go to X Mission hospital. The goats had a combined value of around R1200 but we sold them at R1000 because I wanted to get money to go to hospital. Spiwe_F*<br>*I sold kitchen chairs because I wanted to buy food. At that time I really wanted food, so I sold my kitchen chairs. I sold them for USD25. The market price around that time was USD 120 but I didn't have a choice. Talent_F*<br>*I sold my TV set for USD50 so that I could get money for food. I sold property. At that time it was supposed to fetch at least USD90. Agnella_F.* |
|  | Relocation in search of a caregiver, family support and to reduce cost of transport. | *I went to stay with my mother, the reason being I was looking for someone to assist me, someone who... because I couldn't do anything for myself. I was bathed, sometimes I defecated on myself so I was monitored there. I wanted a caregiver to be around me every time. Talent_F*<br>*There was a time I decided to relocate from... I realized that I had run out of money for transport costs from X to Y. By then I was able get up and walk slowly on my own. Then I finished my course of tablets at the clinic that is in the suburb where I currently live. Talent_F* |
|  | Support from social networks. | *My neighbours would bring me food. In some cases, they would remind me that it was almost time to get to the clinic. I would thank them and tell them that I had fallen asleep. Then I would go [to the clinic].Talent_F* |
|  | Spending savings and defaulting debt payments. | *She explained about my illness to the rotating savings and credit group and fortunately they all know me and they agreed that she be excused in making contributions since she was faced with a problem. They said she had to solve the problem at her home and she would pay back the money when she gets back on her feet. So she was given that lump sum of R11, 000. Up to now she hasn't paid it back. Caleb_M* |
|  | Reduced spending on children's education. | *My daughter failed to sit for her exams that year. She sat for her exams the year that followed. She passed but we could not afford fees for her to continue with education because life was very tough. Even now, life is still tough. Agnella_F* |

### Delays in diagnosis and starting DR-TB treatment

Ten of the 16 participants experienced delays of up to four months before they received a diagnosis and started DR-TB treatment. Reasons for delays included misdiagnosis, HIV co-infection and health seeking from alternative health providers such as traditional healers.

#### Health system diagnostic delays

Reasons for delays in diagnosing DR-TB were multifactorial including the use of tests with low sensitivity (i.e., smear microscopy), lack of healthcare worker awareness and limited health system resources. The use of smear microscopy instead of Xpert MTB/Rif resulted in underdiagnosis of TB and DR-TB. Smear microscopy has a lower sensitivity than Xpert MTB/Rif, especially in people living with HIV, and additionally cannot detect rifampicin resistance.

> *That’s when I went back to the same clinic and to the same nurses who had counselled me and I told them that I had not gained weight and they said, ‘Maybe you have TB. Let’s test you for TB’. When the results came out…they told me that I was TB negative. I was shocked to hear that I did not have TB. They asked me if I was taking my ARVs correctly. Ruth_F*

Though Ruth was finally diagnosed with TB on the basis of smear microscopy, she was initially started on first line TB treatment. Similarly, Godfrey who had had TB in the past and according to the National Guidelines, should have been prioritised for Xpert MTB/Rif testing, was diagnosed using smear microscopy. Hence, he was started on first line treatment. While admitted in hospital, he benefitted from a doctor visiting the facility who requested an Xpert MTB/Rif test. Godfrey felt that he had wasted time on the wrong treatment.

> *He [the doctor] said, ‘I am not talking about the one [smear test] that is done at this hospital. You have to send a sputum sample to the reference laboratory’. They said they hadn’t. He then said, ‘This hospital does not have capacity to do some conclusive tests. What you are doing here are mere jokes’. Godfrey_M*

> *…I had to forget about the previous two months of admission in hospital. It was a joke. [Laughs]. Godfrey_M*

Even where Xpert MTB/Rif was available as a primary diagnostic test, Spiwe was wrongly started on first line TB treatment because healthcare workers misinterpreted the Xpert MTB/Rif result.

> *I took it [first line TB treatment] from November, December to January. Then when I had gone to collect my re-supply in February they told me that I was taking a wrong treatment and was supposed to be on DR-TB treatment. Spiwe_F*

Diagnosing DR-TB in people who have extrapulmonary TB requires invasive sampling. The equipment, consumables and expertise is not readily available in primary health care or district hospitals. Four participants with extra-pulmonary TB experienced delays in diagnosis and initiating appropriate treatment. All four were hospitalised before definite diagnoses were made and required costly laboratory tests and medical procedures that were outsourced to private facilities, as reflected in the experience of Talent.

> *… the injection [needle] they used was thick and long. They inserted it into my stomach and extracted some fluid from there. They took it to a private laboratory and that is how they discovered that I had TB in my stomach. Talent_F*

Five participants experienced delays in turnaround time for follow-up laboratory results, especially during the COVID-19 pandemic. Changes from intensive to continuation phase regimens were only done once follow-up results were received. Longer intensive phase meant that participants were exposed to more drugs and side effects.

> *…*.*so it [long processing times] delays you again from getting your results even though they always come late because of COVID. Felistus_F*

#### Poor health seeking behaviour

Diagnosis of TB was also delayed among participants who first sought help from traditional healers or private clinics. Financial resources for such participants were depleted by the time they were referred to public health facilities. Caleb’s experience shows how some traditional healers would refer people to hospitals.

> *We were also advised by traditional healers to consult hospitals and other traditional healers. They mostly advised us to go to hospitals. That’s when we started going to X [a private clinic]. Caleb_M*

> *We had spent all the money we had by the time I was referred to the doctors at X [TB referral centre] who confirmed that it was TB. Moses_M*

By contrast, those who had had a previous TB episode accessed care earlier at public health facilities, since they were quick to recognise ‘possible TB’ symptoms. Such participants incurred little or no costs and were more likely to be physically fit when they started treatment.

> *I experienced chest pains and night sweats for less than a month. When I started having these symptoms, I said to myself, ‘Maybe the TB that previously infected me was not fully treated. Let me go to the clinic and get tested’. Then I went for testing and the results came out MDR-TB positive. Felistus_F*

### Health service factors undermining quality of TB care

Once people were started on DR-TB treatment, they reflected on how they endured long waiting times at clinics and lack of privacy during administration of injections.

> *There is need for some form of privacy. I can’t pull down my trousers and get an injection while other nurses are watching from one of the rooms. Peter_M*

Three participants were frustrated by lack of service continuity especially in urban clinics. Participants would meet new nurses at each clinic visit. These became more common during COVID-19 when some facilities were closed for fumigation and nurses redeployed to other facilities or services such as COVID-19 duties.

> *We knew each other very well with the nurse who was there and was supposed to administer my injections the week that followed. But that nurse did not come to work because she had been transferred and there was a new nurse. I had to start explaining to her all over again. Peter_M*

Five participants reported that clinics experienced stock-outs of DR-TB drugs during the COVID-19 pandemic. They incurred additional costs because they were asked to travel to neighbouring clinics in search for medicines.

> *Sometimes I would board two commuter omnibuses to get to the clinic only to be told that the drugs were out of stock. They would then refer me to other clinics to check if the tablets were in stock. Moses_M*

Participants living in urban settings were more likely to afford transportation costs compared to those in rural areas. A woman living in a rural village failed to access clofazimine, which meant that she had to go intermittently (during drug stockouts) without clofazimine. She was very frail although she was taking the medicines (except clofazimine) she managed to access religiously.

> *…I’m not getting all the drugs. There is one which was always in short supply; it is called clofazimine. So I wasn’t getting all the drugs for my treatment. I started getting them later. They gave me a full course last month. All along I was getting on an alternate basis. Spiwe_F*

Talent who was on second-line antiretroviral therapy also experienced stock-outs of abacavir from public clinics resulting in a viral load rebound. She incurred substantial costs because she had to purchase abacavir from private pharmacies.

> *In the second line I had a challenge. I was taking Lopinavir and Abacavir. Abacavir is not available in all places. It’s hard to find. So I had a challenge in that abacavir was always out of stock. Being out of stock meant I had to buy it myself. So I would buy monthly supplies at USD7. So this caused my viral load to go down…to increase tremendously [correcting herself]. Talent_F*

### Physical and socioeconomic impact of DR-TB

The impact of DR-TB on households was exacerbated by treatment delays, income instabilities and additional non-TB shocks. People who delayed DR-TB treatment were too frail to walk and work. They were often hospitalised and had to hire cars to visit health facilities because they could not walk from their house to bus stops and from bus stops to health facilities.

> *I wasn’t able to walk coz of weak joints and shortness of breath. So I had to hire a vehicle every day from July-December at R40 [USD2.50] per day. Tecla_F*

Because of the side effects of DR-TB treatment, one man likened the medicines to ‘bulldozers’. Adherence to DR-TB medicines was difficult because of those side effects. The side effects also affected people’s productivity and ability to work. A participant explained that he had to make a tough choice between adhering to medicines and returning to work.

> *Those ones are like a bulldozer. They hit like a big hammer…[laughs]…That treatment hit me hard and made me very very weak and the injection caused me to sleep the whole day. Godfrey_M*

> *Some of us we are in informal trades. Sometimes you would want to be consistent on [adhere to] your medication but it will force you to stop taking your medication and go for work because you can’t get on the street [informal job] when you have taken your medication. Peter_M*

Households experienced huge losses of income during DR-TB treatment. Losses were related to reductions in productivity due to sickness and productive time lost while frequenting healthcare facilities and when household members could not attend to their work or daily chores because they were looking after a sick person. Loss of income was exacerbated by the informal nature of employment which does not reward and allow for absenteeism.

> *…my wife would support me to walk and would carry me on her back. So there was nothing else we could do for our living…My wife stopped doing piece jobs because she was looking after me. Moses_M*

> *…everything changed as soon as I was diagnosed with TB. You can’t get paid when you are not going to…to work. If you don’t go to work everything will also be on hold. Peter_M.*

The physical, psychological and socioeconomic burden associated with DR-TB caused changes in traditional gender roles and strained marital relationships. This in turn led, in some cases, to abandonment by spouses and/or depression. Three women were abandoned by their husbands and Caleb, a man, who had a supportive wife and was bedridden for a long time contemplated suicide.

> *…..after 8 months into my treatment that’s when my husband ran away from us and I was left alone. Life got tough for me and children. Up to now he hasn’t come back. I am still alone. Agnella_F*

> *… I said to my wife, ‘Umn, I have tried a lot to get healed*.*… I have cried a lot. It’s better for me that I go. May God give me rest; I am in pain!’ Caleb_M*

In addition to the dire socioeconomic situation caused by DR-TB, COVID-19 lockdowns led to closure of businesses and forced households to spend their savings and to sell assets. Lockdowns closed a household business for Caleb and shattered job prospects for Godfrey who had recovered from DR-TB and was about to start a new job as a driver.

> *It took a long time before my wife re-opened her kitchen [canteen]. During lockdown, especially the first lockdown, people were not allowed to move that’s when our father sold his car. Life was tough. Caleb_M*

> *Actually we depend on menial jobs. So as of now I…I was thinking of… what’s the name of those companies that are advertised in..? Those earth moving [machines]. So I was ready to go there unfortunately the COVID pandemic forced everything to a standstill. Godfrey_M*

Loss of income was aggravated when participants were abandoned by their breadwinner spouses. Even post DR-TB treatment job opportunities were narrowed with limited prospects to recuperate to their pre-treatment income levels. Ruth who used to earn her livelihood as a hair stylist and was abandoned by her husband felt anxious to start working again.

> *It affected my marriage and my work. I was supposed to be busy plaiting hair right now. If I decide to plait hair, one of the clients may have dandruffs, and with this TB, I may get sick again. So it really affected me. Ruth_29F*

Rural households experienced additional shocks such as drought and pestilences. Pestilences, namely bird flu and tickborne diseases (theileriosis), killed poultry and cattle and amplified the negative effects of DR-TB on household livelihood.

> *That time our chickens got affected by bird flu. We used to rely on chickens for our relish…if you get sick you don’t want to eat substandard foods. Moses_M*

Children were psychologically affected by seeing their parents suffer and in pain. Also they feared their parents might die. Talent explained that education of her child was affected in two ways when the child refused to go to school and when the family could not afford school fees due to depleted financial resources.

> *‘What if I come back from school and find out that my mother is dead’ E-eh right now she was supposed to be in Form 2 but she is in Grade 7 because she refused to go to school. The other ones dropped out of school because as their mother I was not fit enough to run around looking for jobs and assistance. Talent_F*

### Coping strategies adopted by people and their households

Coping strategies were categorised into either reversible or irreversible. All households adopted short term reversible coping strategies such as reducing number of meals and spending savings. Six people relocated to their families in search of support and/or caregivers.

> *It’s like family support. Whenever I took my medication I’d get hungry and I was supposed to cook. I was supposed to…You know? I couldn’t do all of this. So I realized that it was better for me to go back home for maternal care and support. Peter_M*

Support was often provided by immediate families.

> *…my father supported me a lot because that is when some of his pension came out. All of it was spent on my treatment. Loveness_F*

Talent disclosed her DR-TB status to neighbours who assisted her with food and reminders to go to clinic to pick up her medications.

> *My neighbours would bring me food. In some cases, they would remind me that it was almost time to get to the clinic. I would thank them and tell them that I had fallen asleep. Then I would go [to the clinic].Talent_F*

Households that either did not have savings or had spent their savings at the start of treatment initially sold non-productive and then productive assets to raise money for food and transport.

> *We sold chickens to get money to buy sugar, cooking oil or rice. That’s how we survived till we reached a point when she sold our spanners. She’d look around the house to find out if there was something else to sell so that we could get money for bus fare. Moses_M*

Caleb’s livelihood as a private DJ was severely affected following the distress sale of his sound system. He deeply regretted the sale. He coped by making claims on family members to avert further sales of assets.

> *…I ended up selling assets that I wasn’t prepared to dispose. That PA (sound system) system! I am now thinking of replacing the speakers that I sold*

> *Caleb_M*

> *Uum. We realised that we had burdened people. So my wife ended up phoning my mother in-law. My mother-in-law would ask my father-in-law in Harare to send us some money for groceries. That helped us a lot. Caleb_M*

Some households managed to register their children on a government educational social assistance programme. Those who failed to register their children tried to keep their children in school by negotiating with school authorities.

> *In terms of school fees BEAM [Basic Education Assistance Module] came to my rescue. I used to worry and to depend on my mother to pay school fees for my child…Tecla_F*

> *We wanted to register them on BEAM but it seems one needs to have connections in order to get registered. Each time they were sent away from school for not paying fees, my wife would immediately go to the school to plead for their readmission. Moses_M*

## Discussion

Our study revealed huge socioeconomic, physical and psychological impacts of DR-TB among people and their household members, including children. DR-TB pushes households into a downward spiral of poverty and debt traps in a context such as Zimbabwe that has little safety nets and is wracked with economic challenges. Stock-outs of DR-TB drugs were frequently experienced during COVID-19. COVID-19 lockdowns affected people’s livelihoods as people depleted their savings. The convergence of COVID-19, economic challenges and other shocks such as pestilences amplified the impact of DR-TB on households, accelerating irreversible coping strategies.

Diagnostic delays were a common theme in this study. Those diagnosed more rapidly had less severe disease at the time of treatment initiation and incurred less costs. Delays in starting TB treatment have been described in several settings.(42,43) People delay care-seeking because they perceive the quality of services in public clinics as poor or believe that their symptoms are due to other causes.(44) In Zimbabwe, time between onset of symptoms and start of DR-TB treatment was reported to be up to four months as people often sought care from private pharmacies and clinics first before they were diagnosed and referred to public clinics for treatment.(42) Our study shows that health system related factors contribute significantly to these delays.(43) Delayed diagnosis increases the likelihood of physical debility,(45) community transmission of DR-TB, hospitalisation and income loss due to reduced productive capacity by sick people or when households members assume carer roles.(46) As expected, longer diagnostic journeys stretched household resources for a long time such that financial resources were depleted at commencement of DR-TB treatment.(47) DR-TB may break households resilience and may strain social relations with family, neighbours or spouses leading to abandonment, also documented in our study.(45) The End TB strategy aims to reduce TB morbidity and mortality by introducing rapid diagnostics to reduce the time to effective treatment.(18) The full benefits of such technology may not be realised unless delays and structural barriers within the health system are addressed.(42,44)

TB has long-term impacts on income and employment. DR-TB affected households are economically vulnerable even post TB treatment.(6) DR-TB narrows job opportunities and increases likelihood of post TB disabilities such as chronic lung disease. The disabilities result in households continuing to incur health care related costs.(7,8,48,49) In the same way, chronic disabilities as a result of TB or other diseases, recurring TB or additional shocks e.g. COVID-19 and pestilences may break household resilience to shocks faster, rendering households vulnerable to current and future TB.(35)

The socioeconomic situation of DR-TB affected households, already strained due to their disease and related costs, was worsened by COVID-19. Studies have shown that households respond by utilising all resources available to them,(27,28,50) and as observed in this study, may be forced to adopt irreversible coping strategies which had extensive impact on their livelihoods. As observed elsewhere, the impact of DR-TB, combined with COVID-19 was greatest on households without regular income and were food insecure, as they depleted their assets and exhausted short term coping strategies such as borrowing as they were no longer creditworthy.(16,50,51) In the context of distress sales, households may sell more assets to raise the amount of cash they need as assets are usually sold below their market values.(35,50) Though successful treatment outcomes were attained by all participants in this study, this was at the expense of their livelihoods.

Effective TB treatment requires uninterrupted supplies of medicines. Participants reported drug stock-outs during the first few months of the COVID-19 pandemic. This forced them to seek medicines in neighbouring clinics or go without certain medicines. Stockouts of drugs and diagnostics during COVID-19 occurred globally and were enhanced by transport and travel interruptions.(52) The National TB programme decided to dispense three-month supplies of medicines to reduce people’s contact with health facilities.(53) In the context of global supply chain disruptions and increased drug dispensing, stock-outs were therefore inevitable.

This study was conducted between the first (March-July 2020) and second (December-February 2021) COVID-19 lockdowns, reducing the likelihood of recall bias. Most participants included in the study were initiated on DR-TB treatment during the first lockdown which allowed us to understand the challenges at the start of the pandemic. By including participants from rural and urban settings we were able to identify experiences and coping strategies which were setting specific. Participants who were interviewed were either at the end of treatment or had completed treatment providing insights into their diagnostic and treatment journeys. However, by interviewing participants at the end of successful treatment we introduced survival bias.

Therefore, the challenges and coping strategies described here may not be representative of those doing less well on treatment. Studies have reported that DR-TB inflicts huge physical, socioeconomic and psychological impact on household members.(12) However, we only managed to infer the impact of DR-TB on household members based on what the participants affected by DR-TB reported since we did not interview any household members, spouses or children.

Our study has several implications for policy and practice. Firstly, since TB diagnosis is the entry point into treatment and care, delays in diagnosis should be addressed by fostering collaborations between private and public healthcare sectors and alternative health providers such as traditional healers (42,54), investing resources into earlier diagnosis and raising community awareness about signs and symptoms of TB and benefits of early health seeking from public facilities. Secondly, the impact of DR-TB on physical, mental, socioeconomic wellbeing of people and households affected by DR-TB calls for multisectoral approaches even beyond the treatment period, focusing on medical and social services for post TB care and measures to mitigate re-infection with DR-TB.(36)

There are various social support systems provided by government agencies and non-governmental organisations in Zimbabwe including the USD25 cash disbursement every month for the duration of treatment for people on DR-TB treatment. However, evidence from this study and the patient cost survey conducted in Zimbabwe shows that this amount is unlikely to mitigate the costs and income loss experienced by DR-TB households.(14) Importantly, the coverage of the cash scheme is suboptimal and delays and inconsistencies in disbursements are frequent.(38) Additional safety nets in the form of educational assistance; payment of medical bills and extra food and money during and possibly post DR-TB treatment are therefore required. The Ministry of Public Service Labour and Social Welfare manages the Basic Education Assistance Module (for educational social assistance); the Harmonised Social Cash Transfers (for vulnerable households) and the Assisted Treatment Medical Order facility (for formal workers who experience work related disabilities). However, the Assisted Treatment Medical Order does not cover chronic conditions e.g. HIV and TB.(55) Several non-governmental organisations are providing various forms of assistance to vulnerable households within the communities in which we collected data, but their reach is chequered, and coordination between programs is limited and sometimes non-existent. Effective stakeholder mapping and inter-ministerial collaborations aimed at providing social protection to people and households with DR-TB should be included in the National Strategic Plan. People with DR-TB will need to be linked to available social protection. Given that many participants and households included in this study had experienced an episode of TB before and were vulnerable to future DR-TB, social protection should take a broader focus and should be availed before people and households are affected by DR-TB. Such TB sensitive approaches,(56) that focus on food insecure or vulnerable households may have greater impact on TB epidemiology and control in Zimbabwe and will likely prevent future TB/DR-TB episodes.

## Data Availability

All data produced in the present work are contained in the manuscript.

## Conclusion

DR-TB pushes households into poverty, and increases their vulnerability to shocks. Other stressors such as COVID-19, divorce and pestilences enhanced their vulnerability resulting in irreversible coping strategies. The impact of DR-TB extends beyond the duration of DR-TB treatment. Multisectoral approaches combing health system, psychosocial and economic interventions are crucial to reduce diagnostic delays and mitigate physical and mental consequences and loss of livelihoods due to DR-TB and other shocks.

## Acknowledgements

We would like to thank the participants who took part in this study.

## Author contributions

Conceptualised the study: CT, KK, VB.

Reviewed the draft manuscript: CT, KK, DP, VB

Data collection: CT, FM

Data analysis: CT, VB, KK

Critical review of manuscript: CT, KK, FM,VB, FK,SK, DP.

All authors reviewed and approved the final manuscript.

## Data availability statement

The transcripts used in this study have been shared as Supporting information (Transcripts_Coping with DR-TB and COVID-19).

